# Clinical characteristics of pregnant women infected with Coronavirus Disease 2019 in China: a nationwide case-control study

**DOI:** 10.1101/2021.10.21.21265313

**Authors:** Qin Li, Lian Chen, Hai Jiang, Danni Zheng, Yuanyuan Wang, Jie Mei, Xudong Ma, Yuan Wei, Yangyu Zhao, Jie Qiao

**Affiliations:** National Clinical Research Center for Obstetrical and Gynecology, National Center for Healthcare Quality Management in Obstetrics, Ministry of Education Key Laboratory of Assisted Reproduction, Department of Obstetrics and Gynecology, Peking University Third Hospital, Beijing, China; Department of Obstetrics and Gynecology, Sichuan Province People’s Hospital, Chengdu, China; National Health Commission of the People’s Republic China, Beijing, China

**Keywords:** COVID-19, SARS-CoV-2, pregnancy, case-control study

## Abstract

**OBJECTIVE:** To formally compare the clinical course of Coronavirus disease 2019 (COVID-19) in pregnant women with their nonpregnant counterparts.

**METHODS:** Clinical data of pregnant women with confirmed COVID-19 in the designated hospitals of mainland China were retrieved up to April 12, 2020 through an epidemic reporting system maintained at the National Health Commission of the People’s Republic China. Each pregnant patient was randomly matched to a nonpregnant woman with confirmed COVID-19 in the same hospital as control, then their clinical courses were formally compared.

**RESULTS:** 138 pregnant women had been identified as confirmed COVID-19 cases. Among them, 17 severe cases and 1 maternal death were recorded, which was less than their nonpregnant peers (23 severe cases and 3 death). 57.2% had been infected with SARS-CoV-2 during the third trimester, including 13 severe cases and 1 maternal death. 7.3% of pregnant patients had diarrhea and 3.6% had nausea or vomiting, compared with related proportion as 15.2% (OR: 0.38, 95%CI: 0.15, 0.96) and 10.1% (OR: 0.25, 95%CI: 0.07, 0.89) in nonpregnant patients. Pregnant patients infected with SARS-CoV-2 in early pregnancy presented similar laboratory tests with their nonpregnant peers, however, with pregnancy progresses, increased inflammation, coagulation and hepatic injury markers happened more and more frequently (*p*<0.001) in pregnant patients.

**CONCLUSIONS:** Being pregnant did not represent a risk for severe condition when compared with their nonpregnant peers. Patients infected with SARS-CoV-2 in early pregnancy were even at lower risk of severe illness than those infected in late pregnancy.

**What are the novel findings of this work?:** Compared with non-pregnant COVID-19 patients, pregnant patients tend to present less symptom, had unique laboratory findings, and tend to at lower risk of COVID-19–related death. Patients infected with SARS-CoV-2 in the early pregnancy tend to be in the less severe condition of illness than those infected in late pregnancy.

**What are the clinical implications of this work?:** Vital comparisons of the clinical course upon COVID-19 between pregnant and nonpregnant women in childbearing age are, unfortunately, lacking. Through formally comparisons between the two groups, the present study provides more reliable evidence towards the management of pregnant women with COVID-19.

## Introduction

Due to the high infectivity of severe acute respiratory syndrome coronavirus 2 (SARS-CoV-2), coronavirus disease 2019 (COVID-19) caused by the virus has spread quickly in worldwide, and was declared by the World Health Organization (WHO) as a Public Health Emergency of International Concern (PHEIC). Over the past six months, more than 10 million confirmed cases of COVID-19 had been documented globally, resulting in more than 500,000 deaths.^1^

According to the previous studies which were relevant to severe acute respiratory syndrome (SARS) and middle east respiratory syndrome (MERS), pregnant women were more susceptible to coronavirus infection, and they were also at a high risk of poor outcomes, including need for mechanical ventilation, admission to an intensive care unit (ICU), renal failure and death.^2^ Considering that SARS-CoV-2 has up to 80% sequence similarity with SARS-CoV,^3^ a number of literatures suggested that pregnant women might be at higher risk to develop a more severe form of COVID-19 results from the physiological immunosuppression.^4^ More worrying, according to a recently report of the Centre for Disease Control and Prevention of United States (U.S. CDC), 8,207 pregnant women had laboratory-confirmed SARS-CoV-2 infections were documented in America up to June 16, 2020, and the number kept sustainable growing.^5^ Thus, the impact of COVID-19 on pregnant women has drawn great public attention.

Studies include ours tried to describe the clinical features of pregnant COVID-19 patients,^6 7^ while the interaction between SARS-CoV-2 infection and physiological change during pregnancy is not already clear. Vital comparisons of the clinical course upon COVID-19 between pregnant and nonpregnant women in childbearing age are, unfortunately, lacking. It is therefore an emerging need to figure out the questions including whether pregnant women with COVID-19 present distinct manifestations as well as laboratory tests to their nonpregnant peers, whether pregnant women infected with SARS-CoV-2 in different gestation had similar presentation and whether being pregnant represents a risk for severity clinical presentation.

Based on an epidemic reporting system maintained at the National Health Commission of the People’s Republic China (NHCPRC), we conducted a 1:1 case-control study to formally compare the clinical course of COVID-19 in pregnant women with their nonpregnant peers. With the significant results, we are aiming to provide a update insight into the management for pregnant population during the COVID-19 pandemic.

## Methods

### Study design

Patients with COVID-19 in mainland China were admitted in designated hospitals during the outbreak. All designated hospitals were invited to upload the medical records of COVID-19 patients through an epidemic reporting system maintained at NHCPRC. Under the coordination of NHCPRC, we conducted a 1:1 matched case-control study based on the epidemic reporting system. The study was approved in accordance with the agreed procedure with the Ethics Committee of Peking University Third Hospital, waiving written informed consent for deidentified patient data.

### Selection of cases and controls

The aim of the present study was to estimate the impact of pregnant status on the progression of COVID-19. Thus, ‘cases’ were defined as pregnant COVID-19 patients. According to the criterion of Chinese Clinical Guidance for COVID-19 Pneumonia Diagnosis and Treatment (Seventh Edition, released by the NHCPRC), we collected all 138 pregnant women who were confirmed cases of COVID-19 through the epidemic reporting system, including all the 84 laboratory-confirmed cases in Wuhan city which was mentioned in our previous report.^6^ Each pregnant patient was matched to a confirmed patient who was non pregnant, with same age and admitted in same hospital as ‘control’ by simple randomization. 11 cases cannot be matched within the same hospital due to the rare number of COVID-19 patients there, then adequate controls were compensated from other hospitals within the same city. All uploaded medical records of them were retrieved from the epidemic reporting system, then we extracted information regarding epidemiological, clinical, laboratory and radiological characteristics, treatment, and outcomes of them with a customized data collection form. All data collection forms were reviewed by two experienced doctors independently to insure data reliability. Major disagreement between them was resolved by consultation to a third doctor. The data cutoff for the study was from December 8, 2019 to April 12, 2020.

### Study definitions

A confirmed case of COVID-19 was defined as a suspected case with positive result for SARS-CoV-2 on high throughput sequencing or real-time reverse transcriptase polymerase chain reaction (RT-PCR) assay of nasal and pharyngeal swab specimens. The degree of severity (severe and critical case) was classified by using the mentioned guidance. Severe cases were defined as meeting one of these conditions: 1) respiratory rate ≥30 breaths/min; 2) arterial oxygen tension (PaO_2_) over inspiratory oxygen fraction (FIO_2_) of less than 300 mmHg; 3) Percutaneous oxygen saturation (SpO_2_) ≤93% on room air at rest. Critical cases were defined as on the following conditions: 1) occurrence of severe respiratory distress; 2) respiratory failure requiring mechanical ventilation; 3) shock and organ failure even needs of ICU care.

### Statistical analysis

Continuous variables were expressed as medians and interquartile ranges (IQR). Categorical variables were expressed as counts and percentages (%). No imputation was made for missing data. Pairs chi-square test and odds ratios with corresponding 95% confidence interval was used to justify the difference between pregnant patients and nonpregnant patients for categorical features.^8^ Paired t-test was used to justify the difference between pregnant patients and nonpregnant patients for continuous features. We used *p* <0.05 (2-tailed) to determine statistical significance. All statistics analyses were done with R software (R core team, version 3.4.0).

## Results

We identified a total of 138 pregnant COVID-19 patients across mainland China. The largest number of pregnant patients (102) came from Hubei province, followed by the provinces bordering Hubei (Figure 1). Baselines of the pregnant patients and their nonpregnant peers are shown in Table 1. The median age of the pregnant patients was 30.4 years (standard deviation: 4.3). 79 of 138 (57.2%) had been infected with SARS-CoV-2 in the third trimester. A total of 121 pregnant patients (87.7%) had mild disease, while 17 pregnant patients (12.3%) had severe disease (hypoxemia). 8 pregnant patients (5.8%) had complications (hypertension, diabetes, choric hepatitis, or choric nephritis). 1 maternal death was observed in the electronic medical records. For their nonpregnant peers, 23 patients (16.7%) had severe disease, 17 patients (12.3%) had complications (hypertension, diabetes, choric hepatitis, or choric nephritis), and 3 death were recorded.

**Table 1.** Baseline characteristics of pregnant Covid-19 patients and matched comparators.^*^

| **Characteristic** | **Pregnant**<br>**patients**<br>**(n=138)** | **Non-pregnant**<br>**patients**<br>**(n=138)** | ***p-value*** |
| --- | --- | --- | --- |
| **Mean of age (SD)-years** | 30.4 (4.3) | 30.4 (4.3) | 0.857 |
| **Region-no. (%)** |  |  |  |
| Hubei province | 102 (73.9) | 102 (73.9) | 1.000 |
| Other provinces | 36 (26.1) | 36 (26.1) |  |
| **Pregnancy period when infected-no. (%)** |  |  |  |
| First trimester | 28 (20.3) | - | - |
| Second trimester | 31 (22.5) | - |  |
| Third trimester | 79 (57.2) | - |  |
| **Disease Severity-no. (%)** |  |  |  |
| Mild case | 121 (87.7) | 115 (83.3) | 0.306 |
| Severe case | 17 (12.3) | 23 (16.7) |  |
| **Complications-no. (%)** |  |  |  |
| No | 130 (94.2) | 121 (87.7) | 0.048 |
| Yes | 8 (5.8) | 17 (12.3) |  |
| **Outcomes-no. (%)** |  |  |  |
| Cured | 137 (99.3) | 135 (97.8) | 0.341 |
| Death | 1 (0.7) | 3 (2.2) |  |
| **Mean of hospital stay (SD)-days** | 15.9 (8.8) | 15.9 (9.9) | 0.999 |

\* Percentages may not total 100 because of rounding. Covid-19 denotes coronavirus disease 2019, and SD denotes standard deviation.

**Figure 1.**
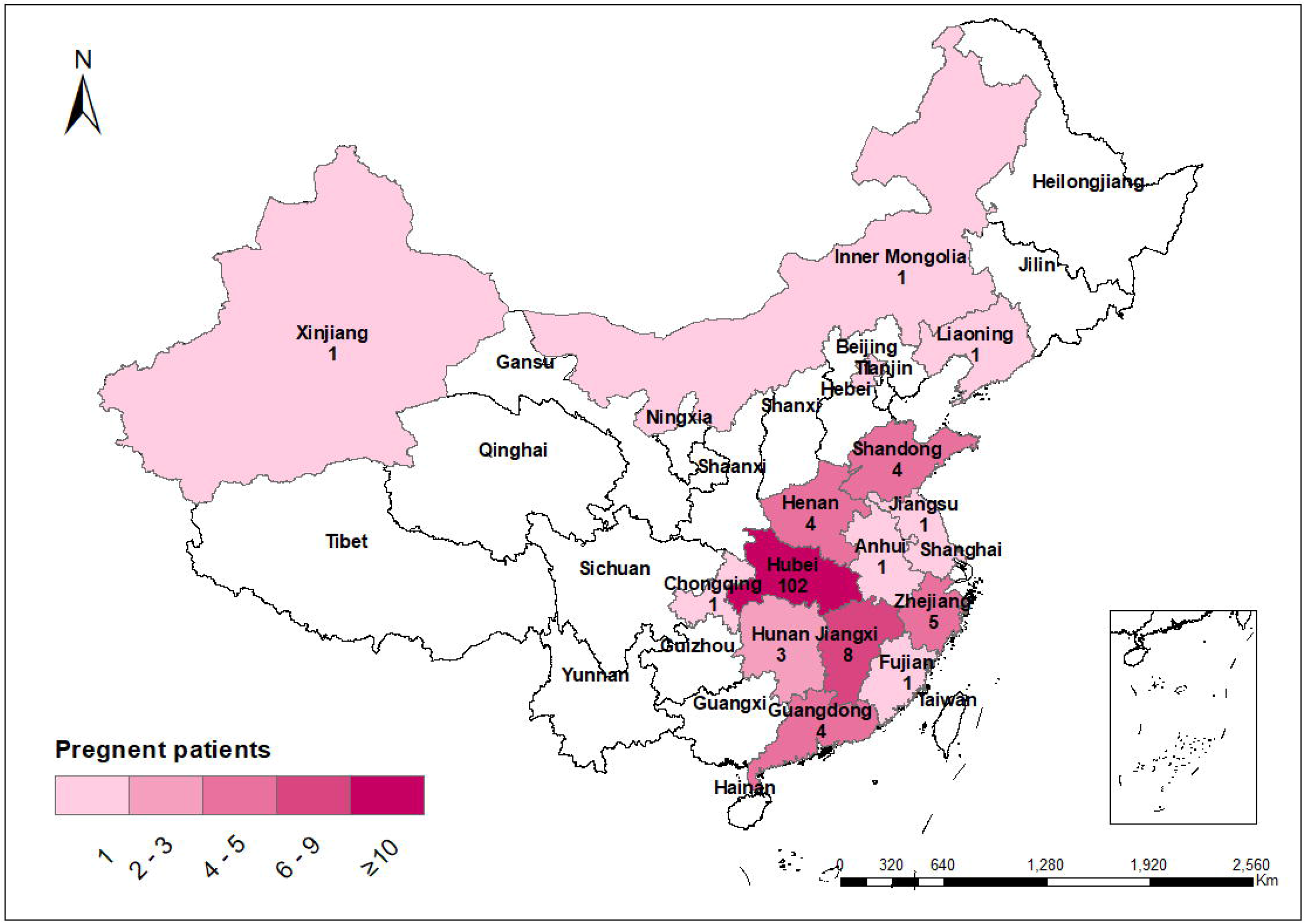
Distribution of pregnant patients with Covid-19 across mainland China. Shown are the uploaded, confirmed pregnant cases of coronavirus disease 2019 (Covid-19) throughout China, according to the National Health Commission as of April 30, 2020.

Table 2 presents the clinical features of pregnant COVID-19 patients by gestation. For manifestations, dyspnea was presented in 15% of the pregnant patients infected with SARS-CoV-2 in the third trimester and presented in 7% of the pregnant patients infected in early pregnancy. For laboratory tests, compared with patients infected with SARS-CoV-2 in early pregnancy, pregnant patient infected in late pregnancy were more likely to present reduced lymphocytes, hemoglobin, and albumin levels (*p*<0.05), as well as elevated fibrinogen, D-dimer, lactate dehydrogenase (LDH), creatine kinase (CK), C-reactive protein (CRP) and procalcitonin (PCT) levels (p<0.05). For treatment, a higher proportion of pregnant patients infected in late pregnancy needed oxygen support (p<0.05) and invasive mechanical ventilation. Overall, we also noticed that 13 of the 17 severe cases were infected in the third trimester (one of them had died), and the other 4 severe cases were infected in the second trimester.

**Table 2.** Clinical features of Covid-19 patients in pregnant.^*^

| Characteristic | Trimester 1<br>(n=28) | Trimester 2<br>(n=31) | Trimester 3<br>(n=79) | p-value |
| --- | --- | --- | --- | --- |
| **Demography** |  |  |  |  |
| Mean of age (SD)-years | 30.7 (5.0) | 30.8 (4.6) | 30.2 (3.9) | 0.692 |
| Living in Hubei Province | 19 (68) | 20 (65) | 63 (80) | 0.171 |
| **Condition-no. (%)** |  |  |  |  |
| Severe case | 0 (0) | 4 (13) | 13 (16) | 0.053 |
| Death | 0 (0) | 0 (0) | 1 (1) | 1.000 |
| **Presenting signs and symptoms--no./total no. (%)** |  |  |  |  |
| Asymptomatic | 1/28 (4) | 1/31 (3) | 9/79 (11) | 0.345 |
| Fever | 18/27 (67) | 21/31 (68) | 56/79 (71) | 0.904 |
| Cough | 17/28 (64) | 28/31 (90) | 50/79 (63) | 0.011<sup>†</sup> |
| Chest tightness | 4/27 (15) | 9/31 (29) | 21/79 (27) | 0.384 |
| Dyspnea | 2/27 (7) | 2/31 (7) | 12/79 (15) | 0.439 |
| Fatigue | 5/27 (19) | 7/31 (23) | 10/79 (13) | 0.367 |
| Diarrhea | 3/27 (11) | 1/31 (3) | 6/79 (8) | 0.535 |
| Myalgia | 6/27 (22) | 2/31 (7) | 4/79 (5) | 0.032<sup>†</sup> |
| Headache | 3/27 (11) | 3/31 (10) | 5/79 (6) | 0.628 |
| Nasal congestion | 1/27 (4) | 4/31 (13) | 5/79 (6) | 0.389 |
| Sore throat | 2/27 (7) | 1/30 (3) | 6/79 (8) | 0.800 |
| Nausea or vomiting | 2/27 (7) | 2/31 (7) | 1/79 (1) | 0.126 |
| Chest pain | 1/27 (4) | 2/31 (7) | 2/77 (3) | 0.589 |
| **Laboratory findings-no./total no. (%)** |  |  |  |  |
| Leucocytes ($\times 10^9$ per mm<sup>3</sup>) | | | | |
| Median (IQR) | 5.9 (5.1-7.5) | 6.9 (4.8-9.3) | 6.6 (5.1-8.8) | 0.254 |
| <3.5×10<sup>9</sup> per mm<sup>3</sup> | 4/28 (14) | 1/31 (3) | 2/76 (3) | 0.078 |
| >9.5×10<sup>9</sup> per mm<sup>3</sup> | 2/28 (7) | 8/31 (26) | 16/76 (21) |  |
| Lymphocytes (×10<sup>9</sup> per mm<sup>3</sup>) |  |  |  |  |
| Median (IQR) | 1.4 (1.1-1.7) | 1.1 (0.8-1.5) | 1.0 (0.8-1.5) | 0.060 |
| <1.1×10<sup>9</sup> per mm<sup>3</sup> | 8/28 (29) | 16/30 (53) | 43/74 (58) | 0.026<sup>†</sup> |
| Hemoglobin (g/liter) |  |  |  |  |
| Median (IQR) | 124 (112-131) | 111 (104-115) | 114 (104-124) | <0.001 |
| Hemoglobin <110 g/liter | 3/25 (11) | 14/30 (47) | 29/74 (39) | 0.005<sup>†</sup> |
| Platelets (×10<sup>9</sup> per mm<sup>3</sup>) |  |  |  |  |
| Median (IQR) | 211 (174-251) | 208 (175-252) | 182 (151-251) | 0.192 |
| <100×10<sup>9</sup> per mm<sup>3</sup> | 0/28 (0) | 0/30 (0) | 3/73 (4) | 0.583 |
| Albumin (g/liter) |  |  |  |  |
| Median (IQR) | 41.8 (38.1-45.3) | 34.4 (31.6-36.4) | 33.2 (31.3-34.7) | <0.001 |
| <35 g/liter | 3/26 (12) | 13/25 (52) | 43/56 (77) | <0.001 |
| Fibrinogen (g/liter) |  |  |  |  |
| Median (IQR) | 3.0 (2.8-4.1) | 4.0 (3.4-5.1) | 4.2 (3.8-4.8) | 0.098 |
| >4 g/liter | 7/25 (28) | 13/25 (52) | 37/58 (64) | 0.011<sup>†</sup> |
| D-dimer (mg/liter) |  |  |  |  |
| Median (IQR) | 0.4 (0.2-1.1) | 0.9 (0.6-1.3) | 1.3 (0.9-2.2) | 0.014<sup>†</sup> |
| ≥0.5mg/liter | 12/26 (46) | 18/23 (78) | 61/64 (95) | <0.001 |
| Alanine aminotransferase (U/liter) |  |  |  |  |
| Median (IQR) | 17 (10-32) | 17 (11-24) | 18 (12-48) | 0.245 |
| >40U/liter | 4/26 (15) | 3/23 (13) | 17/61 (28) | 0.271 |
| >120U/liter | 0/26 (0) | 1/23 (4) | 4/61 (7) | 0.503 |
| Aspartate aminotransferase (U/liter) |  |  |  |  |
| Median (IQR) | 18 (15-24) | 23 (16-28) | 26 (19-43) | 0.113 |
| >40U/liter | 3/26 (12) | 2/23 (9) | 17/60 (28) | 0.071 |
| >120U/liter | 0/26 (0) | 0/23 (0) | 6/60 (10) | 0.109 |
| Lactate dehydrogenase (U/liter) |  |  |  |  |
| Median (IQR) | 161 (146-192) | 179 (159-230) | 207 (180-296) | 0.060 |
| >245U/liter | 3/24 (13) | 5/22 (23) | 19/47 (40) | 0.042<sup>†</sup> |
| Creatine Kinase (U/liter) |  |  |  |  |
| Median (IQR) | 45 (31-49) | 29 (23-39) | 43 (27-105) | 0.021<sup>†</sup> |
| ≥200 U/liter | 0/25 (0) | 0/21 (0) | 4/41 (10) | 0.186 |
| C-reactive protein >10mg/lite | 7/26 (27) | 20/30 (67) | 52/69 (75) | <0.001<sup>†</sup> |
| Procalcitonin ≥0.1ng/ml | 0/20 (0.0) | 2/19 (11) | 15/47 (32) | 0.003 |
| **Radiographic findings-no./total no. (%)** |  |  |  |  |
| Bilateral involvement | 14/23 (61) | 17/20 (85) | 59/75 (79) | 0.144 |
| Unilateral involvement | 6/23 (26) | 3/20 (15) | 14/75 (19) | 0.667 |
| No abnormality | 3/23 (13) | 0/20 (0) | 2/75 (3) | 0.104 |
| **Treatment-no./total no. (%)** |  |  |  |  |
| Antibiotic therapy | 16/28 (57) | 17/31 (55) | 63/79 (80) | 0.010<sup>†</sup> |
| Antiviral therapy | 21/28 (75) | 24/31 (77) | 57/79 (72) | 0.898 |
| Glucocorticoids | 0/28 (0) | 10/31 (32) | 23/79 (32) | <0.001<sup>†</sup> |
| Immune globulin | 2/28 (7) | 7/31 (23) | 16/79 (20) | 0.228 |
| Traditional Chinese medicine | 15/28 (54) | 15/31 (48) | 28/79 (35) | 0.169 |
| Oxygen support‡ | 5/28 (18) | 16/31 (52) | 48/79 (61) | <0.001<sup>†</sup> |
| Mechanical ventilation | 0/28(0) | 0/31 (0) | 3/79 (4) | 0.576 |

\* Percentages may not total 100 because of rounding. Covid-19 denotes coronavirus disease 2019, SD denotes standard deviation, and IQR denotes interquartile range.
<sup>†</sup> p<0.05.
‡ Oxygen support including nasal cannula, high flow oxygen and Invasive mechanical ventilation.

Clinical features of the pregnant COVID-19 patients were compared with their nonpregnant peers in Table 3. Pregnant patients tend to present less symptom than nonpregnant patients during hospitalization. Among the pregnant patients, 16.1% had fatigue, 7.3% had diarrhea and 3.6% nausea or vomiting, however, for their nonpregnant peers, 30.4% had fatigue (OR: 0.44, 95%CI: 0.25, 0.80), 15.2% had diarrhea (OR: 0.38, 95%CI: 0.15, 0.96) and 10.1% had nausea or vomiting (OR: 0.25, 95%CI: 0.07, 0.89). On the other hand, 8.0% of the pregnant patients had no signs or symptoms of the disease, while the proportion was 4.3% among their nonpregnant peers, with a OR of 1.83 (95%CI: 0.68, 4.96).

**Table 3.** Clinical features of Covid-19 patients in pregnant and matched comparator.^*^

| Characteristic | Pregnant patients<br>(n=138) | Non-pregnant patients<br>(n=138) | Odds Ratio (95% CI) or<br>P value for difference |
| --- | --- | --- | --- |
| **Manifestations-no./total no. (%)** |  |  |  |
| Asymptomatic | 11/138 (8.0) | 6/138 (4.3) | 1.83 (0.68 to 4.96) |
| Fever | 95/137 (69.3) | 100/138 (72.5) | 0.87 (0.52 to 1.46) |
| Cough | 96/138 (69.6) | 101/137 (73.2) | 0.83 (0.49 to 1.42) |
| Chest tightness | 34/137 (24.8) | 50/138 (36.2) | 0.61 (0.36 to 1.02) |
| Dyspnea | 16/137 (11.7) | 22/138 (15.9) | 0.71 (0.37 to 1.39) |
| Fatigue | 22/137 (16.1) | 42/138 (30.4) | 0.44 (0.25 to 0.80)<sup>†</sup> |
| Diarrhea | 10/137 (7.3) | 21/138 (15.2) | 0.38 (0.15 to 0.96)<sup>†</sup> |
| Myalgia | 12/137 (8.8) | 22/138 (15.9) | 0.50 (0.23 to 1.07) |
| Headache | 11/137 (8.0) | 21/138 (15.2) | 0.50 (0.22 to 1.11) |
| Nasal congestion | 10/137 (7.3) | 14/138 (10.1) | 0.60 (0.22 to 1.65) |
| Sore throat | 9/137 (6.6) | 15/138 (10.9) | 0.57 (0.24 to 1.36) |
| Nausea or vomiting | 5/137 (3.6) | 14/138 (10.1) | 0.25 (0.07 to 0.89)<sup>†</sup> |
| Chest pain | 5/137 (3.6) | 10/138 (7.2) | 0.50 (0.17 to 1.46) |
| **Laboratory findings-no./total no. (%)** |  |  |  |
| Leucocytes ($\times 10^9$ per mm<sup>3</sup>) | | | |
| Median (IQR) | 6.5 (5.0-8.5) | 4.7 (3.8-6.1) | $p < 0.001$ |
| $< 3.5 \times 10^9$ per mm<sup>3</sup> | 7/135 (5.2) | 24/130 (18.5) | 0.26 (0.11 to 0.64)<sup>†</sup> |
| $> 9.5 \times 10^9$ per mm<sup>3</sup> | 26/135 (19.3) | 3/130 (2.3) | 12.50 (2.96 to 52.77)<sup>†</sup> |
| Lymphocytes ($\times 10^9$ per mm<sup>3</sup>) | | | |
| Median (IQR) | 1.1 (0.8-1.5) | 1.4 (1.0-1.9) | $p < 0.001$ |
| $< 1.1 \times 10^9$ per mm<sup>3</sup> | 67/132 (50.8) | 40/130 (30.8) | 2.04 (1.24 to 3.36)<sup>†</sup> |
| Hemoglobin (g/liter) |  |  |  |
| Median (IQR) | 113 (107-124) | 124 (116-132) | $p<0.001$ |
| <110 g/liter | 46/132 (34.8) | 18/131 (13.7) | 3.64 (1.87 to 7.09)<sup>†</sup> |
| Platelets ($\times 10^9$ per mm<sup>3</sup>) | | | |
| Median (IQR) | 195 (155-244) | 204 (162-252) | $p=0.891$ |
| <100$\times 10^9$ per mm<sup>3</sup> | 3/131 (2.3) | 7/131 (5.3) | 0.33 (0.07 to 1.65) |
| Albumin (g/liter) |  |  |  |
| Median (IQR) | 34.1 (31.6-38.2) | 41.7 (39.2-44.2) | $p<0.001$ |
| <35 g/liter | 59/107 (55.1) | 7/112 (6.2) | 12.75 (4.61 to 35.28)<sup>†</sup> |
| Fibrinogen (g/liter) |  |  |  |
| Median (IQR) | 4.1 (3.3-4.7) | 3.1 (2.5-3.6) | $p<0.001$ |
| >4 g/liter | 57/108 (52.8) | 18/116 (15.5) | 44.00 (6.06 to 319.37)<sup>†</sup> |
| D-dimer (mg/liter) |  |  |  |
| Median (IQR) | 1.1 (0.6-1.8) | 0.2 (0.2-0.4) | $p=0.003$ |
| $\geq 0.5$mg/liter | 91/113 (80.5) | 23/118 (19.5) | 11.00 (4.77 to 25.37)<sup>†</sup> |
| Alanine aminotransferase (U/liter) |  |  |  |
| Median (IQR) | 18 (12-35) | 15 (11-23) | $p=0.003$ |
| > 40U/liter | 24/110 (21.8) | 7/110 (6.4) | 3.17 (1.26 to 7.93)<sup>†</sup> |
| >120U/liter | 5/110 (4.5) | 0/110 (0.0) | Not available |
| Aspartate aminotransferase (U/liter) |  |  |  |
| Median (IQR) | 23 (17-34) | 18 (15-23) | $p=0.005$ |
| > 40U/liter | 22/109 (20.2) | 3/110 (2.7) | 6.00 (1.77 to 20.37)<sup>†</sup> |
| >120U/liter | 6/109 (5.5) | 0/110 (0.0) | Not available |
| Lactate dehydrogenase (U/liter) |  |  |  |
| Median (IQR) | 195 (162-264) | 172 (143-206) | $p=0.089$ |
| >245U/liter | 27/93 (29.0) | 14/99 (14.1) | 2.37 (1.04 to 5.43)<sup>†</sup> |
| Creatine Kinase (U/liter) |  |  |  |
| Median (IQR) | 39 (26-64) | 56 (42-70) | $p=0.320$ |
| $\geq 200$ U/liter | 4/87 (4.6) | 4/88 (4.5) | 0.50 (0.09 to 2.73) |
| C-reactive protein $>10$mg/liter | 79/125 (63.2) | 40/108 (37.0) | 2.77 (1.47 to 5.22)<sup>†</sup> |
| Procalcitonin $\geq 0.1$ng/ml | 17/86 (19.8) | 10/94 (10.6) | 1.33 (0.56 to 3.16) |
| ***Radiographic findings-no./total no. (%)*** |  |  |  |
| Chest CT scan |  |  |  |
| Bilateral involvement | 90/118 (76.3) | 80/131 (61.1) | 2.38 (1.25 to 4.56)<sup>†</sup> |
| Unilateral involvement | 23/118 (19.5) | 38/131 (29.0) | 0.48 (0.24 to 0.96) |
| No abnormality | 5/118 (4.2) | 13/131 (9.9) | 0.50 (0.17 to 1.46) |
| ***Treatment-no. (%)*** |  |  |  |
| Antibiotic therapy | 96 (69.6) | 92 (66.7) | 1.15 (0.68 to 1.95) |
| Antiviral therapy | 102 (73.9) | 132 (95.7) | 0.09 (0.03 to 0.30)<sup>†</sup> |
| Glucocorticoids | 35 (25.4) | 28 (20.3) | 1.33 (0.76 to 2.35) |
| Immune globulin | 25 (18.1) | 14 (10.1) | 2.10 (0.99 to 4.46) |
| Traditional Chinese medicine | 58 (42.0) | 90 (65.2) | 0.36 (0.21 to 0.62)<sup>†</sup> |
| Oxygen support‡ | 69 (50.0) | 59 (42.8) | 1.40 (0.84 to 2.34) |
| Mechanical ventilation | 3 (2.2) | 3 (2.2) | 1.00 (0.20 to 4.95) |

\* Percentages may not total 100 because of rounding. Covid-19 denotes coronavirus disease 2019, and IQR denotes interquartile range.
† $p < 0.05$ .
‡ Oxygen support including nasal cannula, high flow oxygen and Invasive mechanical ventilation.

On admission, the median value of leukocyte counts was 6.5×10^9^/L among pregnant patients, which was higher than that of nonpregnant patients (4.7×10^9^/L, *p*<0.001). 5.2% of the pregnant patients had leukopenia, while 18.5% of the nonpregnant patients had leukopenia, with a OR of 0.26 (95%CI: 0.11, 0.64). The median value of lymphocytes was 1.1×10^9^/L among pregnant patients, which was lower than that of their nonpregnant peers (1.4×10^9^/L, *p*<0.001). 50.8% of the pregnant patients had lymphopenia, while only 30.8% of the nonpregnant patients had lymphopenia, with a OR of 2.04 (95%CI: 1.24, 3.36). When we focus on the women infected in early pregnancy and their nonpregnant peers, the distinctions on leukocytes and lymphocytes disappeared (Table S1, S2 and S3 in appendix). We also found pregnant patients had a lower level of hemoglobin and albumin than their nonpregnant peers (*p*<0.001), but they had a higher level of fibrinogen, D-dimer, alanine aminotransferase (ALT), aspartate aminotransferase (AST), LDH and CRP than their nonpregnant peers (*p*<0.05). Similarly, there was no longer a significant difference on those biomarkers when we focus on the women infected in early pregnancy and their nonpregnant peers (Table S1, S2 and S3 in appendix).

For radiographic findings, 118 of the pregnant patients and 131 of the nonpregnant patients had performed CT scans. 76.3% of the pregnant patients showed bilateral involvement on the chest CT, which was higher than that of their nonpregnant peers (61.1%), with a OR of 2.38 (95%CI: 1.25, 4.56). For treatment, 73.9% of the pregnant patients were given antiviral therapy, which was lower than the proportion of their nonpregnant peers, with a OR of 0.09 (95%CI: 0.03, 0.30). Similarly, the proportion of pregnant patients who received traditional Chinese medicine treatment was 42.0%, which was lower than that of nonpregnant patients, with a OR of 0.36 (95%CI: 0.21, 0.62).

## Discussion

Through the epidemic reporting system maintained at the NHCPRC, we recruited a total of 138 pregnant women confirmed with COVID-19 across mainland China. By analyzing their clinical characteristics, we found pregnant women tend to present less symptom, had unique laboratory findings, and tend to at lower risk of COVID-19–related death. Additionally, we also found patients infected with SARS-CoV-2 in the early pregnancy tend to be in the less severe condition of illness than those infected in late pregnancy. To our best knowledge, this is the first study provide detailed information regarding SARS-CoV-2 infection in different gestation as well as a formally comparison of the clinical course of COVID-19 between pregnant women and nonpregnant women. These findings are essential for understanding of COVID-19 and improving clinical guidance for management of pregnant COVID-19 patients.

In our study, we found the overall incidence of clinical symptoms during hospitalization is lower than that of nonpregnant women, which is consistent with the largest epidemic report of 3,474 pregnant COVID-19 patient from U.S. CDC^5^, as they also indicated that pregnant women less frequently reported headache, muscle aches, fever, chills, and diarrhea than did nonpregnant women. We further noted that the incidence of gastrointestinal symptoms was significantly lower in pregnant women than nonpregnant women, and it was also lower than that of the report of the U.S. CDC (3.6% versus 19.6% had nausea or vomiting; 7.3% versus 14.3% had diarrhea).^5^ The widely use of antiviral drugs may affect the incidence of gastrointestinal symptoms such as nausea and vomiting, thus we had defined symptoms as the incidence at admission to exclude the influence of this factor in the analysis, which may contribute to the difference. Digestive symptoms imply that viral load and replication within the gastrointestinal tract, which was associated with more severe illness as studies had shown that COVID-19 patients without digestive symptoms are more likely to be cured and discharged than patients with digestive symptoms.^9^ These findings suggested that signs and symptoms of COVID-19 might differ between pregnant and nonpregnant women, which can also serve as evidence for the inference that pregnant women tend to have mild illness than their nonpregnant peers.

With the physiological change during pregnancy, many laboratory indicators altered gradually. Comparing to nonpregnant women, the serum albumin and hemoglobin concentration decreases due to hemodilution; coagulation is augmented following with an increased fibrinogen and D-dimer concentration in pregnant population.^10^ Leukocyte counts increased and lymphocytes counts deceased as gestation proceeds due to the gradual increase in neutrophils counts.^11^ When compared pregnant patients with their nonpregnent peers, consistent features were observed as we found leukopenia is less common while lymphopenia and elevated fibrinogen, D-dimer, LDH and CRP levels are more common in pregnant patients than their nonpregnant peers. However, these distinctions showed corresponding trend as pregnancy physiological changes by the gestational week and were observed during the second and third trimesters mainly, which also supported that these distinctions may be caused by the physiological change during pregnancy mainly. Therefore, the immanent physiological characteristics during pregnency should be fully considered when using laboratory test results (e.g., leukocyte and lymphocytes counts) as basis for diagnosis and evaluation of illnesses among pregnant patients.

In contrast, abnormal liver function was more common among pregnant COVID-19 patients than that of their nonpregnant peers. The incidence of abnormal liver function was up to 23% in pregnant patients, and even remained at 15% in women who had been infected with SARS-CoV-2 in early pregnancy, which can hardly be explained by the physiological changes of pregnancy. Prelimilary studies indicated that SARS-CoV-2 can attack several organs which involves not just lung but also heart and liver.^12^ However, with the available evidence at present, we cannot clarify whether pregnancy could increase the risk of liver damage associated with SARS-Cov-2 infection, but monitoring and evaluation of liver function in pregnant patients should be performed and more attention should be paid.

It is well document that analogous coronaviruses such as SARS-CoV infection can cause severe respiratory disease among pregnant women with a high fatality rate.^13^ However, in the nationwide case-control study, we found being pregnant didn’t represent a risk for severity of clinical presentation and maternal death in SARS-Cov-2 infection, which may be related to the special pathogenic mechanism of SARS-Cov-2 and the unique pregnancy immune status of pregnant women.^14^ In the case of SARS-CoV infection, the immune response of Th1 type is highly activated and lasts for a long time, resulting in a consistently high level of pro-inflammatory cytokine expression, which may be responsible for extensive target organ damage.^15^ For SARS-CoV-2 infection, the intensity and duration of Th2 immune responses were similar with Th1,^13^ which may mitigate pro-inflammatory cytokine expression and the resulting organ damage. During pregnancy, the maternal immune system will make real-time adjustments to resist the invasion of pathogens as well as to tolerate the implantation of fetus and placenta.^16^ This specific change of immune state is believed to be related to the tilt of the immune balance of Th1/Th2 to Th2. Th2 predominant immune balance and the role of Treg cells may play an important role in preventing excessive systemic inflammatory response such as cytokines storms and following life-threatening organ failure.^17^ These features of SARS-CoV-2 infection and pregnancy may contribute to the relatively mild illness among pregnant patients. On the other hand, Little, Zheng and their colleagues suggested that viral load, defined as the size of the infecting dose of SARS-CoV-2, is likely to be important for the severity of disease,^18 19^ which implied that the mild condition of pregnant women may also associated with their lower exposure due to strengthen self-protection measures.

Among pregnant COVID-19 patients, we noted that patients infected with SARS-CoV-2 in the late pregnancy tend to be in the more severe condition of illness than those infected in early pregnancy, as 13 of the all 17 severe cases in our study were infected in the late pregnancy (one of them had passed away), and a higher proportion of pregnant patient infected in late pregnancy presented dyspnea who needed oxygen support or even invasive mechanical ventilation. Higher oxygen demands due to pregnancy-associated anatomical and physiological adaptive changes (e.g., diaphragm elevation, increased oxygen consumption, incremental circulation volume and edema of respiratory tract mucosa) in the cardio-respiratory system, increased risk of pulmonary microvascular thrombosis due to pregnancy-associated hypercoagulable state could all have an important role in the clinical course of SARS-CoV-2 infection during the third trimester of pregnancy.^20^

We also noted that a certain number of maternal deaths caused by COVID-19 were reported by various country and regions.^5 21 22^ According to the report of U.S. CDC, there were 16 pregnant COVID-19 patients had passed away. They also reported that pregnant patients were 1.5 times more likely to be admitted to the ICU (95% CI: 1.2, 1.8) and 1.7 times more likely to receive mechanical ventilation (95% CI: 1.2, 2.4). Another national study from Sweden also found that pregnant patients were 5 times more likely to be admitted to the ICU and 4 times more like to receive mechanical ventilation than were nonpregnant patients.^23^ In our study collecting data up to the April 12, 2020, only 1 maternal death at 35 days after delivery was recorded, which was lower than that of nonpregnant patients. Pregnant patients in our study were also less likely to be in the more severe condition of illness. Even not much was known about the SARS-CoV-2 in early days of the pandemic, the high priority of prenatal care, the effective performance of the maternal and child health care system, the principle of leaving no one unattended, the practice of early diagnose and hospitalization, and the positive supportive treatment under the guide of a multidisciplinary team may contribute to the favorable prognosis of pregnant COVID-19 patients in this study. However, considering the differences of maternal death across regions, further comparisons of the virus strains between different countries may be instructive.

The findings of present study also provide more reliable evidence towards the management of pregnant women with COVID-19. For mild patients infected with SARS-CoV-2 during early pregnancy, as being pregnant did not associate with increased risk of severe illness, and the prognosis for most of them was favorable, we had no evidence sufficient to terminate pregnancy in current stage. However, special attention must be paid to patients infected with SARS-CoV-2 in the late pregnancy since the risk of severe illness among them seems higher than that of patients infected in early stage and a exacerbations of COVID-19 were observed in women during the postpartum period.^6^ While for severe or critical cases, it is necessary to take both obstetrical situation and the severity of the condition into consideration, to terminate pregnancy at the best time to prevent the occurrence of serious adverse maternal and fetal outcomes.

It is also worth noting that the findings of the present study are subject to limitation of missing data. We can hardly rule out the existence of missed case due to the pregnant COVID-19 cases were identified based on the uploaded medical records. However, we undertook a meticulous process in reviewing previous literatures and then believed that the present study had the capability to capture a national picture of the impact of COVID-19 on pregnancy. Additionally, given the variation in the clinical process of diagnosis and treatment and the electronic medical records databases among the designated hospitals, some cases had missing values in the laboratory testing. It may lead to misestimation of some characteristics if those with missing data were systematically different from those with available data. Further, collection and comparation of longitudinal data for pregnant women with and without SARS-CoV-2 infection during the outbreak is needed to understand the effects of SARS-CoV-2 infection on maternal and neonatal outcomes.

In summary, compared with nonpregnant COVID-19 patients, being pregnant did not represent a risk for severe condition. Moreover, patients infected with SARS-CoV-2 in early pregnancy were at lower risk of severe illness than those infected in late pregnancy.

## Data Availability

All data produced in the present work are contained in the manuscript.

## Contributors

QL, LC, HJ and DNZ designed the study, collected the data, interpreted the results, and drafted the manuscript. JM collected the data, interpreted the results, and revised the manuscript. JQ, YYZ, YW, DXM conceived of the study, supervised the study, interpreted the results, and revised the manuscript. QL, LC, HJ and DNZ contributed equally and are considered as co-first authors. JQ, YYZ, YW and DXM contributed equally and are considered as co-corresponding authors. All authors contributed to critical reading of, and commented on, the manuscript, helped to interpret the data, and approved the final manuscript.

## Declaration of interests

The authors declare no conflicts of interest.

## Acknowledgments

We thank Yanhong Guo, Xinqiang Gao, Qiang Wang (National Health Commission of the PRC), for their approval and support on provision and coordination of collecting data; We would also thank Ying Shi, Luxia Gan (Big Data Centre of National Health Commission for Human Tissue, Organ Transplantation and Medicine), Qimin Zhan, Luxia Zhang, Mai Wang, Na Cui and Yang Xing (National Institute of Health Data Science at Peking University) for providing help for collecting data. We thank all the hospital staff members for their efforts in collecting the information that was used in this study, and all the patients who consented to donate their data for analysis and the medical staff members who are on the front line of caring for patients; We would also thank the editor and peer reviewers for their hard work for this article.

## Funding

This study was financially supported by Chinese Academy of Engineering (2020-KYGG-01-06), National Natural Science Foundation of China (72042013), Peking University Health Science Center (BMU2020HKYZX001). The mentioned foundation had no rule in the study design, data analysis, drafting the manuscript, or decision to submit this article for publication.

## Notes

### Competing Interest Statement

The authors have declared no competing interest.

### Funding Statement

This study was funded by Chinese Academy of Engineering (2020-KYGG-01-06), National Natural Science Foundation of China (72042013), Peking University Health Science Center (BMU2020HKYZX001).

### Author Declarations

Ethics committee of Peking University Third Hospital gave ethical approval for this work.

## Reference

1. World Health Organization. Coronavirus disease (COVID-19) Situation Report – 164. Geneva: World Health Organization, 2020.

2. Tang P, Wang J, Song Y. Characteristics and pregnancy outcomes of patients with severe pneumonia complicating pregnancy: a retrospective study of 12 cases and a literature review. BMC Pregnancy Childbirth 2018;18(1):434. doi: 10.1186/s12884-018-2070-0 [published Online First: 2018/11/06]

3. Zhou P, Yang XL, Wang XG, et al. A pneumonia outbreak associated with a new coronavirus of probable bat origin. Nature 2020;579(7798):270–73. doi: 10.1038/s41586-020-2012-7 [published Online First: 2020/02/06]

4. Liu H, Wang LL, Zhao SJ, et al. Why are pregnant women susceptible to COVID-19? An immunological viewpoint. J Reprod Immunol 2020;139:103122. doi: 10.1016/j.jri.2020.103122 [published Online First: 2020/04/04]

5. Ellington S, Strid P, Tong VT, et al. Characteristics of Women of Reproductive Age with Laboratory-Confirmed SARS-CoV-2 Infection by Pregnancy Status - United States, January 22-June 7, 2020. MMWR Morb Mortal Wkly Rep 2020;69(25):769–75. doi: 10.15585/mmwr.mm6925a1 [published Online First: 2020/06/26]

6. Chen L, Li Q, Zheng D, et al. Clinical Characteristics of Pregnant Women with Covid-19 in Wuhan, China. N Engl J Med 2020;382(25):e100. doi: 10.1056/NEJMc2009226 [published Online First: 2020/04/18]

7. Yan J, Guo J, Fan C, et al. Coronavirus disease 2019 in pregnant women: a report based on 116 cases. Am J Obstet Gynecol 2020;223(1):111 e1–11 e14. doi: 10.1016/j.ajog.2020.04.014 [published Online First: 2020/04/27]

8. Pearce N. Analysis of matched case-control studies. BMJ 2016;352:i969. doi: 10.1136/bmj.i969 [published Online First: 2016/02/27]

9. Pan L, Mu M, Yang P, et al. Clinical Characteristics of COVID-19 Patients With Digestive Symptoms in Hubei, China: A Descriptive, Cross-Sectional, Multicenter Study. Am J Gastroenterol 2020;115(5):766–73. doi: 10.14309/ajg.0000000000000620 [published Online First: 2020/04/15]

10. Tan EK, Tan EL. Alterations in physiology and anatomy during pregnancy. Best Pract Res Clin Obstet Gynaecol 2013;27(6):791–802. doi: 10.1016/j.bpobgyn.2013.08.001 [published Online First: 2013/09/10]

11. Naccasha N, Gervasi MT, Chaiworapongsa T, et al. Phenotypic and metabolic characteristics of monocytes and granulocytes in normal pregnancy and maternal infection. Am J Obstet Gynecol 2001;185(5):1118–23. doi: 10.1067/mob.2001.117682 [published Online First: 2001/11/22]

12. Zhang C, Shi L, Wang F-S. Liver injury in COVID-19: management and challenges. The Lancet Gastroenterology & Hepatology 2020;5(5):428–30. doi: 10.1016/s2468-1253(20)30057-1

13. Forestieri S, Marcialis MA, Migliore L, et al. Relationship between pregnancy and coronavirus: what we know. J Matern Fetal Neonatal Med 2020:1–12. doi: 10.1080/14767058.2020.1771692 [published Online First: 2020/06/06]

14. Breslin N, Baptiste C, Gyamfi-Bannerman C, et al. COVID-19 infection among asymptomatic and symptomatic pregnant women: Two weeks of confirmed presentations to an affiliated pair of New York City hospitals. Am J Obstet Gynecol MFM 2020:100118. doi: 10.1016/j.ajogmf.2020.100118 [published Online First: 2020/04/16]

15. Wong CK, Lam CW, Wu AK, et al. Plasma inflammatory cytokines and chemokines in severe acute respiratory syndrome. Clin Exp Immunol 2004;136(1):95–103. doi: 10.1111/j.1365-2249.2004.02415.x [published Online First: 2004/03/20]

16. Aghaeepour N, Ganio EA, McIlwain D, et al. An immune clock of human pregnancy. Science Immunology 2017;2(15) doi: 10.1126/sciimmunol.aan2946

17. Sarapultsev A, Sarapultsev P. Immunological environment shifts during pregnancy may affect the risk of developing severe complications in COVID-19 patients. Am J Reprod Immunol 2020:e13285. doi: 10.1111/aji.13285 [published Online First: 2020/06/10]

18. Little P, Read RC, Amlot R, et al. Reducing risks from coronavirus transmission in the home-the role of viral load. BMJ 2020;369:m1728. doi: 10.1136/bmj.m1728 [published Online First: 2020/05/08]

19. Zheng S, Fan J, Yu F, et al. Viral load dynamics and disease severity in patients infected with SARS-CoV-2 in Zhejiang province, China, January-March 2020: retrospective cohort study. BMJ 2020;369:m1443. doi: 10.1136/bmj.m1443 [published Online First: 2020/04/23]

20. Westgren M, Pettersson K, Hagberg H, et al. Severe maternal morbidity and mortality associated with COVID-19: The risk should not be downplayed. Acta Obstet Gynecol Scand 2020;99(7):815–16. doi: 10.1111/aogs.13900 [published Online First: 2020/05/10]

21. Hantoushzadeh S, Shamshirsaz AA, Aleyasin A, et al. Maternal Death Due to COVID-19 Disease. Am J Obstet Gynecol 2020 doi: 10.1016/j.ajog.2020.04.030 [published Online First: 2020/05/04]

22. Amorim M, Soligo TM, Fonseca E. Maternal deaths with coronavirus disease 2019: a different outcome from low-to middle-resource countries? Am J Obstet Gynecol 2020 doi: 10.1016/j.ajog.2020.04.023

23. Collin J, Byström E, Carnahan A, et al. Pregnant and postpartum women with SARS-CoV-2 infection in intensive care in Sweden. Acta Obstet Gynecol Scand 2020 doi: 10.1111/aogs.13901 %/ This article is protected by copyright. All rights reserved.

